# Protocol for scoping review to Identify and characterize Surgery, Obstetric, Trauma and Anesthesia care in Ugandan health policy databases

**DOI:** 10.1101/2022.09.22.22280231

**Authors:** Kasagga Brian, Berjo Dongmo Takoutsing, Balumuka Darius, Ambangira Fortunate, Nawezo Jacob, Kasozi Derrick, Namiiro Amelia Margaret, Sekyanzi John, Chebet Isaac, Jean Namatovu Kizito, Namazzi Mercy, Yusuf Sadiq, Mutatina Boniface, Ekwaro Obuku, Elobu Alex Emmanuel

## Abstract

**Introduction:** Diseases addressed by surgical, obstetrics, trauma, and anesthesia (SOTA) care are rising globally due to an anticipated rise in the burden of non-communicable diseases and road traffic accidents. Low and middle-income countries (LMICs) disproportionately bear the brunt. Evidence-based policies and political commitment are required to reverse this trend. The Lancet Commission of Global Surgery proposed National Surgical and Obstetrics Plans (NSOAP) to alleviate the respective SOTA burdens in LMICs. NSOAP plans success leverages comprehensive stakeholder engagement and appropriate health policy analyses and recommendations. As Uganda embarks on its NSOAP development, policy prioritization in Uganda remains unexplored. We, therefore, seek to determine the priority given to Surgery, Obstetrics, Anesthesia, and Trauma care in Uganda’s health care policy and systems-relevant documents.

**Methods and analysis:** We will conduct a scoping review of SOTA health policy and system-relevant documents produced between 2000 and 2022 using the Arksey and O’Malley methodological framework and additional guidance from the Joanna Briggs Institute Reviewer’s manual. These documents will be sought from the websites of SOTA stakeholders by hand searching. We shall also search from Google scholar and Pubmed using well-defined search strategies. The Knowledge Management Portal for the Ugandan Ministry of Health, which was created to provide evidence-based decision-making data, is the primary source. The rest of the sources will include; other repositories like websites of relevant government institutions, international and national non-governmental organizations, professional associations and councils, and religious and medical bureaus. Data retrieved from the eligible policy and decision-making documents will include the year of publication, the global surgery specialty mentioned, the NSOAP surgical system domain, the national priority area involved, and funding. The data will be collected in a preformed extraction sheet. Two independent reviewers will screen the collected data, and results will be presented as counts and their respective proportions. The findings will be reported narratively using the PRISMA guidelines for scoping reviews.

**Ethics and dissemination:** This study will generate evidence-based information on the state of SOTA care in Uganda’s health policy, which will inform NSOAP development in this nation. The review’s findings will be presented to the Ministry of Health planning task force. The study will also be disseminated through a peer-reviewed publication, oral and poster presentations at local, regional, national, and international conferences, and over social media.

**Strengths and Limitations of the study:** This will be the first scoping review to examine the prioritization of SOTA care in Uganda’s health care policy documents. The search strategy includes several electronic databases, including governmental and non-governmental organizations, professional associations and councils, and religious and medical bureaus. The scoping review will conform to the rigorous methodology manual by the Joanna Briggs Institute.

However, this scoping review may not capture some documents that aren’t available online.

## BACKGROUND

Eleven percent of the global disease burden is treatable by surgical care^1^; however, approximately five billion people lack access to timely, safe, and affordable surgical, obstetric, trauma, and anesthesia (SOTA) care when needed^2^. This huge disparity exists mainly in low- and middle-income countries (LMICs), where nine out of ten people do not have access to SOTA care^2^.

Until recently, Surgery and anesthesia were considered burdensome, luxurious, and less cost-effective aspects of global health^3^. The Lancet Commission on Global Surgery (LCoGS) in 2015 projected an increased need for SOTA care in LMICs to effectively address the burden of communicable and non-communicable diseases and road traffic accidents. Mindful of these challenges, a new term, “Global surgery,” was adopted to describe a rapidly developing multidisciplinary field aiming to provide improved timely, safe, and affordable SOTA care across international health systems with a focus on LMICS such as Uganda.^4^

This new term brought about numerous academic and policy stimuli. These included landmark publications such as the World Bank’s third edition of their Disease Control Priorities (DCP-3) and World Health Assembly (WHA) resolution 68.15, which was adopted unanimously by the World Health Organisation Member States in 2015. This resolution calls for strengthening emergency and essential surgical and anesthesia services as a part of universal health coverage^5^. The LCoGS, with its published seminal report, focused on workforce, training, education; healthcare delivery and management; information management; and economics and finance for SOTA care and championed the most significant stimuli in the same year ^4^. It proposed six indicators to be monitored, evaluated, and reported by all countries and global health organizations^4^. These efforts ultimately culminated in recommendations to develop National Surgical and Obstetric Plans (NSOAP), using the facility- and country-level data to drive health policy. Since then, governments, ministries, professional societies, and on-the-ground clinicians have been interested in leading efforts to increase surgical, obstetric, and anesthesia care in their countries by developing the National Surgical, Obstetric, and Anaesthesia Plans (NSOAPs)^6,7^.

Recognizing current gaps in its surgical system’s six core health domains, the Zambian Ministry of Health implemented resolution 68.15 by developing an NSOAP in the country at the beginning of 2016. The goal was to integrate the NSOAP plan into the National Health Sector Strategic Plan of Zambia, 2017-2021.^8^ Following this, other sub-Saharan African countries, including Madagascar, Tanzania, Rwanda, Ethiopia, and Nigeria, have developed similar NSOAP plans. Uganda currently has no active NSOAP plan.

Uganda, a low-income country with inadequate health budget allocation and a high burden of out-of-pocket expenditure of approximately 38% (National Health Accounts, 2018/19), faces several challenges in timely access and affordable and safe SOTA care.^9^ Most of the facilities lack surgical amenities to perform bellwether procedures.^10^ The surgical volume in 2011 was estimated at 241 per 100,000, which has not changed over the last ten years.^11^ The mean national specialist surgical workforce density is about 1.05 for every 100,000 people.^12^ Moreover, poor remuneration and difficult working environments have led to poor retention of the surgical workforce within Uganda.^13^ Within an average catchment area of 23.99 km, the mean proportion of Ugandans living 30 minutes, 1 hour, and 2 hours from a surgical facility is 64%, 87%, and 98%, respectively.^14^ Furthermore, Ugandan SOTA patients face significant financial hardship. The risks of catastrophic and impoverishing SOTA-care-related expenditures are estimated at 65% and 69%, respectively.^15,16^ Regarding safety, the maternal mortality ratio was 375 per 100,000 live births in 2017. Also, the neonatal and under-5 mortality rates per 1,000 births were estimated at 19 and 43, respectively, in 2020.^17,18^

In light of these challenges, the Ugandan Ministry of Health in 2020 embarked on developing NSOAP plans in collaboration with various stakeholders.^19^ NSOAPs are designed to strengthen surgical systems, covering every health system domain: infrastructure, surgical workforce, service delivery, information management, governance, and financing in alignment with national health plans. In developing NSOAPs, existing health policies should be considered and analyzed to avoid redundancy and identify opportunities for collaboration, pooling of resources, and synergy. This analysis may also inform the development and review of evidence-based policies. Although there have been some research efforts in other countries to identify opportunities for SOTA (Takoutsing et al. 2021 etc.), no such efforts have focused on SOTA health policy analysis in Uganda. Thus, this study will seek to determine the priority given to Surgery, Obstetrics, Anesthesia, and Trauma care in Uganda’s health care policy and decision-making documents.

### 1.0 STUDY OBJECTIVES

This protocol aims to describe the methodology of an up-to-date scoping review of existing Ugandan national health policies and related documents (strategies, plans, guidelines, rapid response summaries, health events, evidence briefs for policy, and dialogue reports) and identify opportunities for SOTA policies. The primary and secondary aims of the review are described in Box 1. The findings from this study will inform the ongoing NSOAP development and implementation that commenced in 2020 and, as a result, may lead to progress toward adhering to the global Surgery 2030 agenda.^19^

#### Box 1.

**Primary and secondary aims of the review.**

Primary aim:

- To determine the extent to which SOTA is prioritized in national health policy and systems documents.

Secondary aim

- To characterize all documents with SOTA as a focus area in terms of origin, type, year of production, target audience, health system domain covered, and national priority areas.

### 2.0 Methods and Analysis

We shall review all relevant health policy and decision-making documents about the Uganda health system and interventions on all available health databases produced from 2000 to 2022. We chose to commence from 2000 as this year was critical for health sector reforms. Before 1990 Uganda faced political instability. The focus of the new government was on restoring law and order. Funding and resources for social services such as health was limited and came mostly from external donors who focused on specific disease programs in just a few districts. These efforts were unsustainable, and thus health outcomes stagnated between 1990 and 2000. The year 2000 marked the beginning of implementing important health sector reforms in Uganda with a sector-wide approach. These included the National Health Policy and Health Sector Strategic Plan (2000/01-2004/05).^20^ The study design and identification of relevant documents are adopted from previous health policy analysis by Mutatina et al., in which they attempted to establish a one-stop shop for health policy documents in Uganda.^21^

#### Protocol Design

Arksey and O’Malley’s framework informed the design of the proposed scoping review methodology, which in a health policy research context includes five stages to conducting a scoping review: (1) identifying the research question; (2) identifying relevant studies; (3) study selection; (4) charting the data; (5) collating, summarizing and reporting the results. We will also draw insights from the PCC (Population, Concept, Context) framework to modify stage 2 to fit our design involving identifying policy documents. The future corresponding scoping review will be reported according to the Preferred Reporting Items for Systematic Reviews and Meta-Analyses Extension for Scoping Review guidelines.^22,23^

#### Stage 1: Identifying the research question

After consultation with the entire research team, the overall research questions were;

1. Between 2000 and 2022, what proportions of different health policy and systems documents address specific SOTA aspects;-Surgery, obstetrics, trauma, and anesthesia care?
2. What prominence is SOTA given in key policy documents (plans, guidelines, strategies, and policies)
3. What is SOTA care’s volume and nature (type, content areas/scope, year of production, target audience/ stakeholders, and funding) in Uganda’s health policy documents between 2000 and 2022?
4. How are the SOTA-related policies distributed in the six domains of the surgical system; governance, infrastructure, service delivery, workforce, information management, and health financing?
5. What proportion of SOTA-related policies focuses on Bellwether or essential surgical procedures?
6. Did the advent of key Global Surgery stimuli in 2015 and COVID-19 in 2019 cast any changes in SOTA-related Uganda’s decision-making documents?

#### Stage 2: Identifying relevant policy documents

The PCC (Population, Concept, Context) framework proposed by the Joanna Briggs Institute manual is adopted in this review, as demonstrated in Table 1. According to this framework, the population is Ugandans of any age group requiring a surgical procedure. The concept is to prioritize surgical obstetrics, anesthesia, and trauma care, whereas the context is SOTA care in a low-income setting, that is to say, Uganda.

**Table 1.0.** PCC framework for scoping review.

| **PCC element** | **Definition** | **Example** |
| --- | --- | --- |
| Population | Ugandans of any age group needing a surgical procedure. | 1. Pregnant mother who needs Spinal anesthesia for Caesarean section following obstructed labor.<br>2. Neonate with malrotation requiring general anesthesia and surgical correction.<br>3. Casualty of a motorcycle road traffic accident requiring fracture management. |
| Concept | To determine the extent to which SOTA is prioritized in national health policy and systems documents | 1. Searching for SOTA keywords in health policy documents.<br>2. Determining the prominence given to SOTA in key policy documents (plans, policies, strategies, guidelines) |
| Context | Access to SOTA care for those who need it in a low-income country, Uganda. | Health policy and systems arrangements in Uganda that enable access to surgical care, for example;<br>1. Surgical Infrastructure( healthcentres and hospital, Medicines, blood, and equipment)<br>2. Workforce( training and education of SOTA related staff; anesthesia providers, nurses, surgeons, etc) |

##### Databases and search strategy

We conducted a stakeholder mapping of all institutions involved in delivering SOTA care in Uganda(**Table 2.1**). A targeted hand search/manual strategy will be executed on the websites of the above institutions. We shall use the Google search engine to find such websites, which we then navigate using the tabs and menus available on the home page (e.g., policy documents and guidelines, e-library, resources, publications, legislation). Since different websites are organized differently, we shall develop specific search strategies for each website depending on its navigability. In addition, we shall search Google Scholar and Pubmed using the following keywords in various combinations with Boolean operators (and, or); Uganda, health policy, health system, policies, strategies, plans, and reports. (**Table 2.2**). We shall inspect the reference lists of found documents to expand our list of included documents. Importantly, we shall use the websites from stakeholder mapping as an entry point to other repositories for national strategy documents. The above search strategy was adapted from Takoutsing et al. 2022 and Mutatina et al. 2017.

**Table 2.1.**
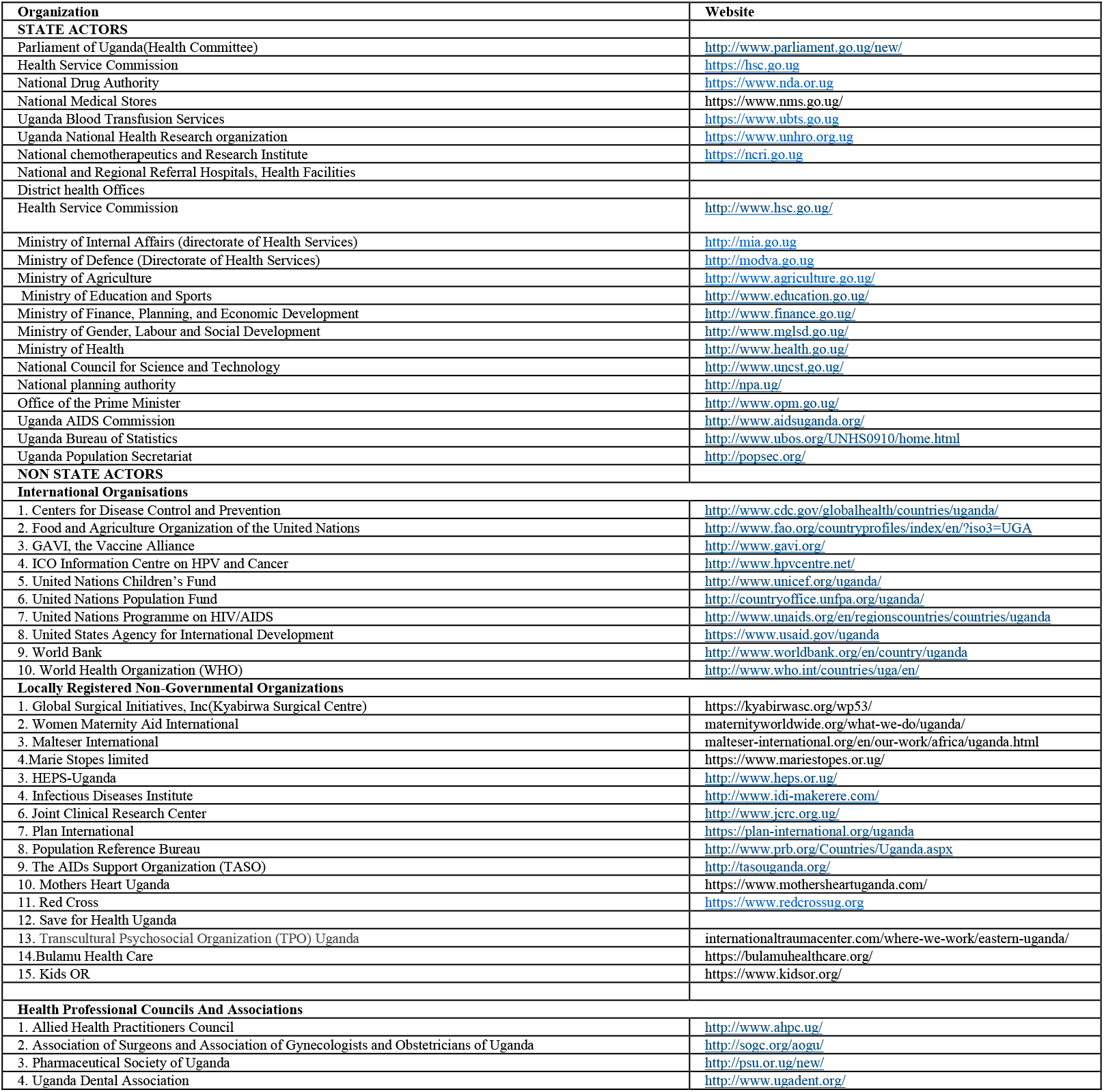
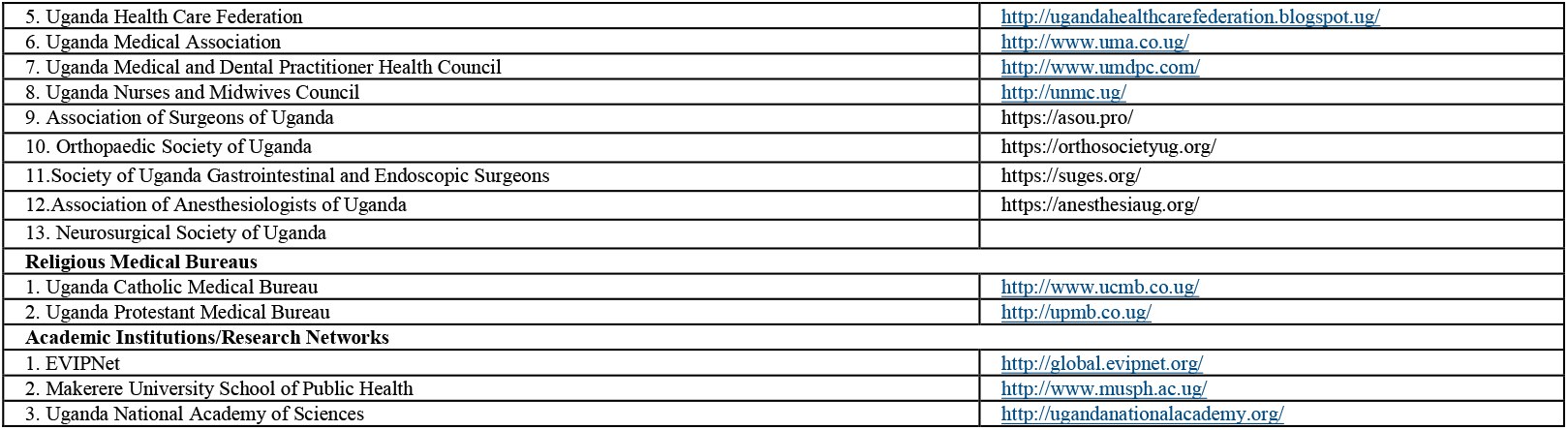
Stakeholders in SOTA care in Uganda.

**Table 2.2.** Pilot search strategy of Google scholar and Pubmed databases.

| PUBMED |  | RESULTS |
| --- | --- | --- |
|  | 1. Surgery OR obstetric* OR trauma OR anesthesia |  |
|  | 2.health polic* OR plan OR strateg* OR rapid response summar* OR guideline OR report* OR evidence brief* OR Roadmap* OR evidence synthes* |  |
|  | 3. 1 AND 2 |  |
|  | 4. Uganda |  |
|  | 5. 3 AND 4 |  |
|  | 6. 2000-2022 |  |
|  | 7. 5 AND 6 | 308 |
| GOOGLE SCHOLAR | (Surgery OR obstetric* OR trauma OR anesthesia) AND (health policy OR plan OR strateg* OR rapid response summar* OR guideline OR report* OR evidence brief* OR Roadmap* OR evidence synthes*) AND Uganda | 121 |

#### Stage 3: Screening and selection of relevant documents

At this stage, KB and BDT will discuss and use the keywords incorporating the inclusion criteria. The search strategy will be developed by testing the keywords, and MESH terms, on the databases to search. Finally, all reviewing team members will discuss and determine the final search strategy.

To delineate the boundaries of SOTA health policy and systems research, we borrowed the Hoffman et al. model to determine the documents relevant to health policy and systems to incorporate in this study.^24^

We shall therefore include all SOTA-related documents between 2000 and 2022 that address the following;

1. issues related to health systems (i.e., SOTA leadership and governance, financial and service delivery, health information systems, and implementation strategies);
2. policies on clinical issues, which include essential drugs, diagnostics, and medical supplies, for example, blood transfusion for surgical operations.
3. policies on public/population issues such as breast cancer screening, and immunization for cancer prevention, for example, HPV vaccine for cervical cancer. We shall exclude documents that;
  1. Have no national coverage (e.g., NGO project reports covering only a few districts).
  2. Are in the draft stage
  3. Are primary studies(case reports, series,reviews,cross-sectional and cohort studies)

The screening and selection will be conducted in three phases. First, all articles identified per our search will be exported to an Excel proforma sheet (Microsoft, Redmond, Washington, USA), and duplicates will be identified and deleted. Next, a calibration exercise will be carried out before screening to ensure an adequate understanding of the inclusion criteria by study screeners. At least two independent reviewers will review each policy document. The documents identified by either or both reviewers will be included for extraction. Disagreements will be discussed amongst the reviewers, and in case of no resolution, an appeal will be made to a senior authors (EO, BM, and EAE). A flow diagram will be presented to reflect the search process.

#### Stage 4: Charting the data

Key characteristics from the included studies will be extracted using a predefined data-extraction sheet in Microsoft Excel (Microsoft, Richmond, Virginia, USA). Data extraction will also be performed in two stages. Firstly a pilot stage consisting of all authors independently extracting and categorizing data from ten policy documents. This is to assure the reliability and standardization of the extraction form and that all authors extract data homogeneously and accurately. Next, the authors will complete data extraction for all included documents, and discrepancies that were not resolved between the authors will be arbitrated by the senior authors (BM, EO, and EAE). The expected key information to be extracted is outlined in Box 2. A sample tabular extraction form is also illustrated in Table 3.

##### Box 2.

**Key information to be extracted.**

- Year of publication of policy document
- Type of document
- Global surgery SOTA aspect focussed on.
- Health system components involved
- Stakeholders involved
- Funding

**Table 2.** Preliminary findings of the SOTA documents

| Doc S/N | Title | Year | Source | Type of document | Global surgery/SOTA aspect | NSOAP surgical domain | Bellwether procedure mentioned | National Priority area | Focus area | Funding |
| --- | --- | --- | --- | --- | --- | --- | --- | --- | --- | --- |
| 1 | National Medicines Policy | 2015 | MoH | Policy | Unspecified | Infrastructure | Unspecified | Essential medicines and supplies | Access to good quality, affordable medicines, and health supplies | MoH |
| 2. | The National Policy Guidelines and Service Standards for Reproductive Health Services | 2001 | MoH | Guidelines | Obstetrics | Service delivery | Cesarean section | Governance arrangements | Streamlining the training and provision of Reproductive Health services | USAID |
| 3. | Improving Access to Skilled Attendance at Delivery | 2012 | SURE project | Evidence brief for policy | Obstetrics | Service Delivery | None | Delivery arrangements | Increasing Access to Skilled Birth Attendance, | European Commission's Seventh Framework Programme |

#### Stage 5: Collating, summarizing, and reporting the results

The SOTA scoping review results will be presented narratively, describing the documents’ scope and nature. The data will be summarised with descriptive statistics in graphs and tables; regarding types of documents, their focus, NSOAP domains, and national priority areas or issues addressed in these documents. We shall also expose the trends, especially during the COVID-19 era and following the publication of key Global Surgery documents in 2015.

## Data Availability

All data produced in the present study are available upon reasonable request to the authors.

## 3.0 Ethics

Ethical approval for this study will not be required because this study did not involve human participants.

## 4.0 Dissemination

This will include: presenting the review’s findings to the Ministry of Health planning task force; publication of the protocol and the review in peer-reviewed journals; oral and poster presentations at local, regional, national, and international conferences; and dissemination over social media.

## 5.0 Competing interests

None declared.

## 6.0 Patient consent for publication

Not required.

## 7.0 Funding

There is no funding for this study

## 8.0 Author’s contributions statement

KB, BDT, and BD were responsible for conceiving the article. KB is the guarantor. KB, BDT, and BD wrote the manuscript. EAE, BD, BM, and EO critically appraised the manuscript. All authors critically revised and approved the final protocol.

## Notes

### Competing Interest Statement

The authors have declared no competing interest.

### Funding Statement

This study did not receive any funding

### Summary of Updates

This version of the manuscript has been revised to update the following Methodology with the PCC framework guided by the Joanna Briggs Manual Stakeholders in SOTA care Search strategy Sample preliminary findings from SOTA documents

